# Pre-existing and new users of psychotherapy and pharmacotherapy for common mental disorders during the COVID-19 pandemic: a population-based study

**DOI:** 10.1101/2025.09.24.25336577

**Authors:** Marie Pouquet, Camille Davisse-Paturet, Cécile Vuillermoz, Marion Debin, Charly Kengne-Kuetche, Clément Turbelin, Thierry Blanchon, Olivier Steichen, Marie Tournier, Nadia Younes

**Author notes:** Author for correspondence: Marie Pouquet, 27 rue de Chaligny 75012 Paris.

## Abstract

**Background:** The COVID-19 pandemic increased mental health service use. The extent of treatment for common mental disorders (CMD) and the profile of new users remain poorly understood. This study aimed to assess the prevalence of psychotherapy and pharmacotherapy use for CMD in the French general population, and to compare the characteristics of new users during the pandemic with those of pre-existing users with respects to need, predisposing and enabling factors of healthcare use.

**Methods:** We conducted a cross-sectional study within the French Grippenet/Covidnet cohort in April 2022. Data were collected through a voluntary online questionnaire. Within the overall sample (n=4,159), weighted analyses estimated the prevalence for psychotherapy or pharmacotherapy use for CMD. Among users of these treatments (n=1,092), logistic regression models compared characteristics of new users during the pandemic versus pre-existing users.

**Results:** Overall, 26.1% (95%CI [24.4-27.8]) of participants reported receiving treatment for CMD: 16.0% [14.5-17.5] psychotherapy, and 18.0% [16.6-19.5] pharmacotherapy (anxiolytics: 12.2%, [11.0-13.4], antidepressants: 8.0%, [7.0-9.0], and hypnotics: 3.6%, [3.1-4.0]). Compared with pre-existing users (n=862), new users (n=230) were more likely to receive psychotherapy alone (45.8% vs. 25.9%, p < 0.0001), and less likely to use pharmacotherapy (54.2% vs. 74.1%, p < 0.0001). Compared with pre-existing users, new users of psychotherapy were more likely to report a decline in mental health during the pandemic (adjusted odd ratio [aOR]=2.63, 1.70-4.12), a history of symptomatic COVID-19 (aOR=1.54, 1.02-2.32), and to use pharmacotherapy (aOR=2.83, 1.68-4.84), but less likely to report financial difficulties (aOR=0.45, 0.23-0.87), or to have used pharmacotherapy before the pandemic (aOR=0.15, 0.08-0.27). New users of pharmacotherapy were more likely to report a decline in mental health during the pandemic (aOR=1.87, 1.28-2.74), to be a female (aOR=1.54, 1.02-2.39), to use psychotherapy (aOR=4.79, 2.87-8.08), but less likely to use psychoactive substances (aOR=0.59, 0.36-0.94) and to have used psychotherapy before the pandemic (aOR=0.14, 0.08-0.25).

**Conclusions:** One in four French adults was receiving treatment for CMD, with new users showing distinct treatment patterns and characteristics, highlighting the importance of addressing pandemic-related mental health needs and reducing financial barriers to psychotherapy.

## 1. Introduction

The COVID-19 pandemic led to an increase in common mental disorders (CMD), including anxiety, depression, and sleep disturbances particularly in high-income regions [1–3]. Although prevalence peaked during the pandemic, recent studies reveal it remains elevated years later, especially among adolescents [4]. Women, younger individuals, those with pre-existing mental or physical health conditions, and those facing financial difficulties were among the most affected groups [5]. Timely and equitable access to evidence-based interventions - ensuring that those in need can obtain them and that gaps in access are identified and addressed - is essential to alleviate symptoms and prevent long-term consequences. This includes psychotherapy for all CMD severity levels according to current recommendations, and psychotropic medications for moderate to severe cases [6].

While increased mental health service use during the pandemic has been widely reported, the extent to which psychotherapy and pharmacotherapy are used for CMD in the general population remains insufficiently described. Disruptions in mental health services early in the pandemic were followed by a rebound in care use, particularly for anxiety and depression [7]. Similar trends were observed in the use of psychotropic medications, including antidepressants and benzodiazepines [8, 9]. Increased service use was especially reported among women and younger individuals [10, 11]. These trends likely reflect increased mental health needs rather than improved access, as evidence suggests that the treatment gap persisted during the pandemic, along with inequities in access to care [10, 12–18].

According to Andersen’s behavioral model of health services use, factors influencing healthcare utilization can be categorized into need factors (perceived or evaluated mental health status), predisposing factors (demographic and social characteristics), and enabling factors (individual and community resources that influence access) [19, 20]. Comparing these factors between new users - those initiating treatment during the pandemic - and pre-existing users can provide insights into changing access patterns, highlighting shifts in needs or potential inequities in access to care. A Canadian study suggested differences in need, predisposing, and enabling factors between new and pre-existing users of mental health services during the pandemic, but didn’t distinguish between one-time users and those receiving ongoing treatment, limiting its ability to assess patterns of new therapy use. [18]. Moreover, it did not distinguish between psychotherapy or pharmacotherapy whereas this distinction is critical, as these treatments have different access pathways: pharmacotherapy is typically prescribed by general practitioners and covered by insurance, while psychotherapy requires more active engagement, including seeking a specialist, attending regular sessions, and often paying out-of-pocket [21–23]. A French study on new psychotropic users during the pandemic focused mainly on age and sex, without considering a broader range of factors [24].

The objectives of this study were to: 1) assess the prevalence of psychotherapy and pharmacotherapy use for CMD in the French general population two years after the onset of the pandemic; 2) compare treatment patterns and characteristics of new users with those of pre-existing users based on Andersen’s model.

## 2. Methods

### 2.1. Study design and participants

We conducted a cross-sectional study within the French Grippenet/Covidnet cohort. This cohort is a participatory online system established in 2012 to monitor acute respiratory infections in the general population [25]. This cohort has previously been utilized in multiple studies addressing diverse health topics, not limited to infectious diseases [26, 27]. All adult participants (≥18 years, n=6,944) were invited by email to complete a survey specific to this study on mental health care and associated factors in April 2022. Because the age structure of the cohort population differed markedly from that of the general French population, analyses were adjusted for age groups (cf. statistical methods) [26].

Demographic, socio-economic status, and chronic non-psychiatric comorbidities were obtained from the annual Grippenet/Covidnet inclusion survey, while mental health-related information were collected through the dedicated questionnaire.

For the analysis of prevalence, all respondents who completed the mental health survey were included. For the comparison of new and pre-existing users, analyses were restricted to participants who reported current use of psychotherapy or pharmacotherapy for CMD at the time of the survey. In both cases, individuals with extreme statistical weights were excluded to ensure robust analyses (see statistical analysis section for details).

Informed consent was obtained from participants during registration. The study was approved by the French Advisory Committee for research on information treatment in the field of health (CCTIRS, authorization 11.565) and the French National Commission on Informatics and Liberty (CNIL, authorization DR-2012-024).

### 2.2. Measures

#### 2.2.1. Psychotherapy and pharmacotherapy use

Psychotherapy for CMD was assessed using the question: “Are you currently receiving regular support from a formal healthcare provider (e.g., psychiatrist, psychologist, general practitioner, or other medical doctor) aimed at improving sleep, addressing emotional problems, or promoting relaxation, excluding the use of medications?“[28, 29].

Pharmacotherapy for CMD was assessed with the question: “Within the past month, have you taken an antidepressant or a medication prescribed for relaxation or sleep?” Participants also provided names of the medications, which were verified and classified as antidepressants, anxiolytics, or hypnotics using the Anatomical Therapeutic Chemical (ATC) classification system [30].

Participants who reported current use of psychotherapy or pharmacotherapy were asked whether they had started the treatment before or during the pandemic, and were classified as ‘pre-existing users’ or ‘new users’ accordingly

#### 2.2.2. Need factors

Need factors included symptoms of CMD. Depressive symptom severity was assessed using the 9-item Patient Health Questionnaire (PHQ-9), a self-report tool evaluating depressive symptom severity in the last two weeks [31]. According to this scale, scores of 0–4 indicate none or minimal depression symptoms, 5–9 mild, 10–14 moderate, 15–19 moderately severe, and 20–27 severe ones. Anxiety symptom severity was measured using the 7-item Generalized Anxiety Disorder scale (GAD-7), which assesses generalized anxiety symptoms in the last two weeks [32]. According to this scale, scores of 0– 4 indicate none or minimal anxiety symptoms, 5–9 mild, 10–14 moderate, and 15–21 severe ones. Insomnia symptom severity was assessed using the 7-item Insomnia Severity Index (ISI), which measures symptoms in the last month [33]. According to this scale, scores of 0–7 indicate absence of insomnia, 8–14 subthreshold insomnia, 15–21 moderate, and 22–28 severe one.

Other need factors included self-rated mental health (good/moderate/poor) [34], chronic non-psychiatric conditions, substance use (including tobacco, cannabidiol, cannabis, cocaine, ecstasy and other drugs, assessed as yes/no), and several pandemic-related factors: perceived negative psychological impact from the pandemic, a history of symptomatic COVID-19, the cumulative COVID-19 hospitalizations number by department (per 100,000 inhabitants, based on government data) as a proxy for the severity of the pandemic, being a public-facing worker (defined as anyone working in direct contact with the public, including healthcare and non-healthcare workers) during the pandemic.

#### 2.2.3. Predisposing factors

Predisposing factors included age, sex, educational level (secondary education or less vs higher education), living with adult(s) (as a proxy for reduced social isolation during the pandemic), history of adverse life events, and experience of a stressful life event within the past 12 months (e.g. divorce, death of a loved one, job loss), assessed with a checklist of specific events to which participants responded “yes” or “no.”

#### 2.2.4. Enabling factors

Enabling factors included 1) individual socio-economic and psychosocial factors: being employed, financial status (good/neither good nor difficult/difficult), three measures of social support (feeling supported financially, feeling supported in daily life, and feeling supported morally or emotionally), loneliness (4-point scale ranging from “Very lonely” to “Very surrounded”) ; 2) three contextual factors: the French ecological deprivation index (based on unemployment rate, labor rate, high school graduation rate, and median household income [35]), the density of general practitioners per 100,000 inhabitants (defined by the French national health insurance, www.ameli.fr), and the urban/rural classification (defined by the French national statistical institute www.insee.fr); 3) behavioral factors, which may alleviate symptoms of CMD, potentially delaying or preventing the need for professional care, particularly in the context of the pandemic: physical activity, relaxation techniques (meditation, breathing exercises, sophrology, others), over-the-counter (OTC) medications to improve sleep, emotional balance, or aid relaxation; 4) one pandemic-related factor: the deterioration of the financial situation during the pandemic (perceived as unchanged, improved, or degraded).

### 2.3. Statistical analysis

Missing data (2.6% all values in the dataset) were handled using multiple imputation by chained equations (MICE) [36].

We first used the overall sample to describe its characteristics as raw counts and weighted percentages, and to estimate the weighted prevalence of psychotherapy and pharmacotherapy use. Weights were used to account for sampling design: post-stratification weights, aligned with age, sex, and regional distributions from the French National Institute of Statistics (INSEE), were multiplied by inverse probability weights derived from logistic regression models (covariates: region, urban/rural residence, age, sex, education, socioeconomic status, household size, smoking, and comorbidities). Final weights were calibrated using generalized raking to match the 2020 population distributions (INSEE). Extreme weights (>99th percentile) were trimmed to reduce outlier influence (n=40 excluded), and weights were normalized to the sample size. Prevalence estimates were reported with 95% confidence intervals (CIs).

Then, we restricted the analysis to current users of psychotherapy or pharmacotherapy, to compare new users with pre-existing users. To examine group differences in prevalence, a Fisher’s exact test was conducted. To identify the most important factors that distinguish new users of psychotherapy and pharmacotherapy during the pandemic from those who were already using these treatments, we used the least absolute shrinkage and selection operator (LASSO) method for feature selection [37, 38]. This method helps simplify the model and reduces issues with multicollinearity by shrinking less important coefficients to zero, leaving only the key predictors. In each model, with either psychotherapy or pharmacotherapy use as the outcome, we tested two interaction terms to assess whether the associations between predictors and the outcome differed according to concurrent use or prior (pre-pandemic) use of the other treatment.

All analyses were conducted in R version 4.0.2, using the ‘mice’ package for weighting and ‘glmnet’ for LASSO. We considered results statistically significant at a 5% level, with two-tailed p-values.

## 3. Results

### 3.1. Overall sample description

The final sample comprised 4,159 participants. Their characteristics (raw counts and weighted percentages) are presented in Supplementary material S1. The median age was 57 years (interquartile range: 40-67; min-max: 18-91), and 54.0% were females. Overall, 25.0% of individuals were living without another adult, 57.3% reported an adverse life event and 47.4% a difficult event in the past 12-months.

Regarding need factors, 23.6% exhibited moderate to severe symptoms of CMD, including depressive symptoms (17.0%), anxiety symptoms (13.5%), and insomnia symptoms (9.1%). Self-rated mental health was reported as moderate by 25.0% of participants and poor by 7.3%. Additionally, 43.0% reported a decline in their mental health during the pandemic.

Regarding enabling factors, 8.2% reported being in a difficult financial situation, 18.0% experienced a financial decline during the pandemic, and 24.3% reported feeling lonely. A lack of moral, daily, or financial support was reported by 10.7%, 18.7%, and 24.3% of participants, respectively.

In terms of behavioral factors, 64.9% engaged in regular physical activity, 25.5% used OTC medications and 45.8% practiced relaxation or mindfulness activities (36.0%: breathing exercises; 17.2%: meditation; 7.3%: yoga or similar practices; 0.5%: alternative medicines).

### 3.2. Psychotherapy and pharmacotherapy prevalence

Overall, 26.1% (95% CI [24.4-27.8]) of the population received treatment for CMDs. Psychotherapy was used by 16.0% (95% CI [14.5-17.5]) of the population, while 18.0% (95% CI [16.6-19.5]) used pharmacotherapy. Regarding treatment types, 8.1% (95% CI [6.9-9.3]) received psychotherapy alone, 10.2% (95% CI [9.1-11.2]) received pharmacotherapy alone, and 7.9% (95% CI [6.9-9.0]) received both psychotherapy and pharmacotherapy. Anxiolytics were the most commonly used psychotropics (12.2%, 95% CI [11.0-13.4]), followed by antidepressants (8.0%, 95% CI [7.0-9.0]) and hypnotics (3.6%, 95% CI [3.1-4.0]).

### 3.3. Comparison of new and pre-existing users

#### 3.3.1. Treatment patterns

Among the 1,092 users of psychotherapy or pharmacotherapy, 21.1% (n = 230) were classified as new users, while 78.9% (n = 862) were pre-existing users. Significant differences in treatment patterns were observed between these groups (Table 1). New users had significantly lower rates of pharmacotherapy use compared to pre-existing users (54.2% vs. 74.1%, p < 10^−3^). This pattern was consistent across all classes of psychotropics, with significantly lower use of antidepressants (19.7% vs. 34.1%, p = 0.02) and anxiolytics (36.9% vs. 49.9%, p = 0.05) among new users. In terms of treatment modalities, new users were more likely to receive psychotherapy alone compared to pre-existing users (45.8% vs. 25.9%, p < 10^−3^). Conversely, combined psychotherapy and pharmacotherapy use was significantly less common among new users (19.5% vs. 33.8%, p = 0.02).

**Table 1.** Prevalence of psychotherapy and pharmacotherapy use for common mental disorders in the general population in France in April 2022, and comparison between new and pre-existing users during the COVID-19 pandemic.

| Treatments for CMDs | Overall sample<br>(n = 4,159) |  |  | New users<br>(n = 230) |  |  | Pre-existing users<br>(n = 862) |  |  | p-value |
| --- | --- | --- | --- | --- | --- | --- | --- | --- | --- | --- |
|  | n<sup>a</sup> | %<sup>b</sup> | 95%CI | n<sup>a</sup> | %<sup>b</sup> | 95%CI | n<sup>a</sup> | %<sup>b</sup> | 95%CI |  |
| Psycho- or pharmacotherapy | 1,092 | 26.1 | (24.4-27.8) |  |  |  |  |  |  |  |
| Psychotherapy | 611 | 16.0 | (14.5-17.5) | 135 | 65.3 | (57.3-72.8) | 476 | 59.7 | (55.7-63.6) | 0.37 |
| Psychotherapy alone | 285 | 8.1 | (6.9-9.3) | 81 | 45.8 | (37.2-54.5) | 204 | 25.9 | (22.1-30.0) | <10<sup>-3</sup> |
| Pharmacotherapy | 807 | 18.0 | (16.6-19.5) | 149 | 54.2 | (45.5-62.8) | 658 | 74.1 | (70.0-77.9) | <10<sup>-3</sup> |
| Pharmacotherapy alone | 481 | 10.2 | (9.1-11.2) | 95 | 34.7 | (27.2-42.7) | 386 | 40.3 | (36.4-44.3) | 0.37 |
| Antidepressants | 344 | 8.0 | (7.0-9.0) | 58 | 19.7 | (14.2-26.1) | 286 | 34.1 | (30.2-38.2) | 0.02 |
| Anxiolytics | 538 | 12.2 | (11.0-13.4) | 100 | 36.9 | (29.2-45.1) | 438 | 49.9 | (45.7-54.1) | 0.05 |
| Hypnotics | 178 | 3.6 | (3.1-4.0) | 32 | 10.8 | (7.0-15.6) | 146 | 14.8 | (12.4-17.5) | 0.31 |
| Psycho- and pharmacotherapy | 326 | 7.9 | (6.9-9.0) | 54 | 19.5 | (13.9-26.1) | 272 | 33.8 | (29.8-37.9) | 0.02 |

<sup>a</sup> Unweighted observed numbers. <sup>b</sup> Weighted percentages. CMD: common mental health disorders.

#### 3.3.2. Psychotherapy users

Table 2 presents a comparison of characteristics between new and pre-existing users of psychotherapy for CMD during the pandemic. Univariate analyses are detailed in Table 2. In multivariate analyses, compared to pre-existing users, new users of psychotherapy were more likely to report a decline in mental health during the pandemic (OR = 2.63, 1.70-4.12) and to have a history of symptomatic COVID-19 (OR = 1.54, 1.02-2.32). New users were less likely to face financial difficulties (OR = 0.45, 0.23-0.87). They were more likely to be concurrent users of pharmacotherapy (OR = 2.83, 1.68-4.84), but less likely to have started it before the pandemic (OR = 0.15, 0.08-0.27).

**Table. 2.**
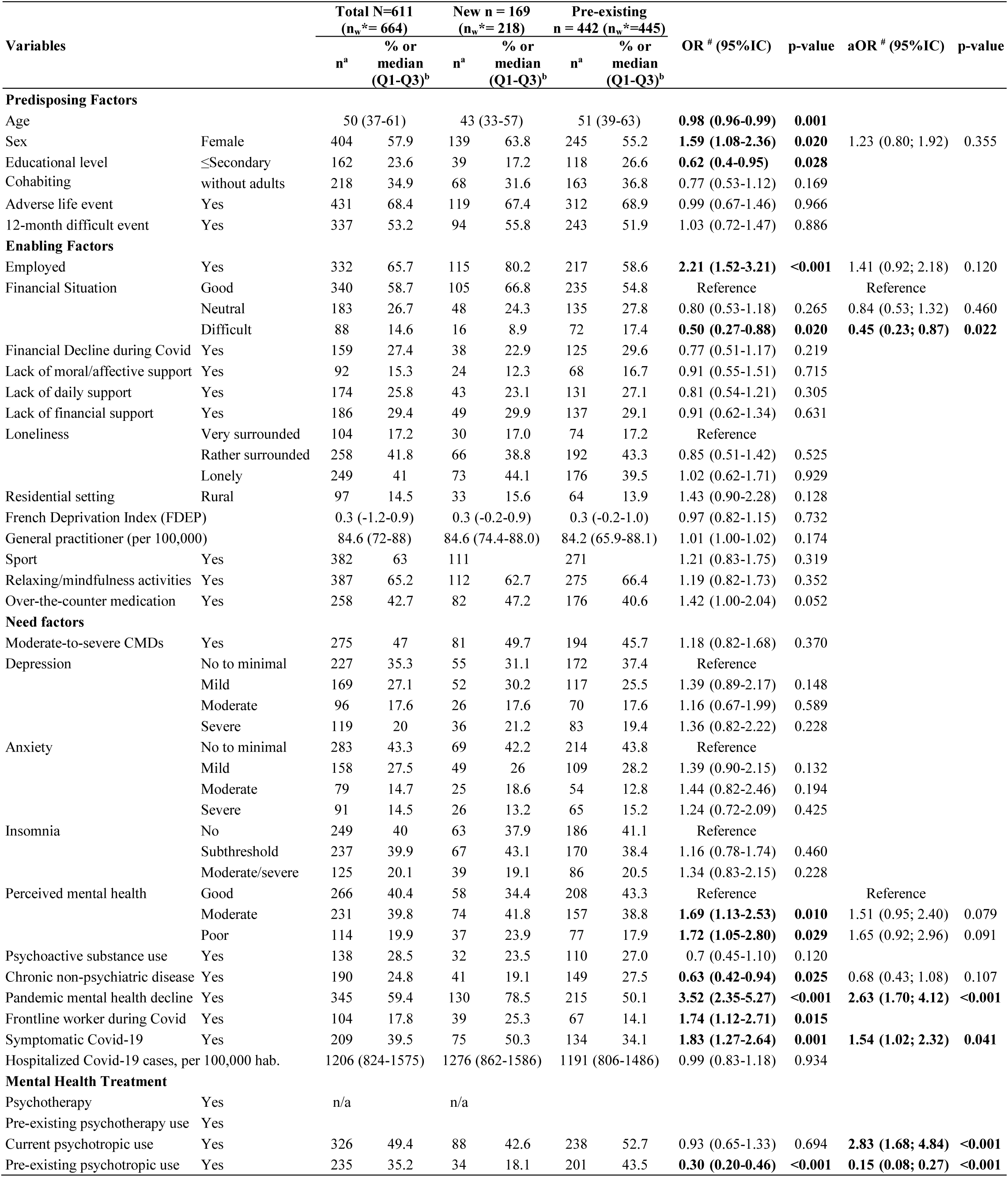
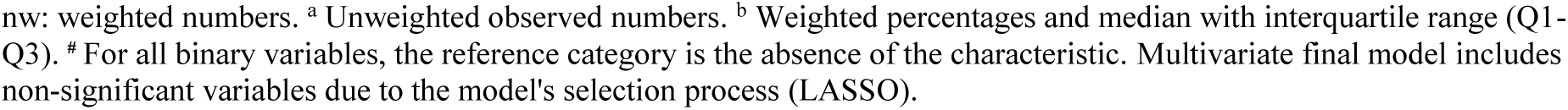
Comparison between new and pre-existing users of psychotherapy for common mental disorders during the pandemic.

#### 3.3.3. Pharmacotherapy users

Table 3 presents a comparison of characteristics between new and pre-existing users of pharmacotherapy for CMD during the pandemic. Univariate analyses are detailed in Table 3. In multivariate analyses, compared to pre-existing users, new users of pharmacotherapy were more likely to report a decline in mental health during the pandemic (OR = 1.87, 1.28-2.74) and to be a female (OR = 1.54, 1.02-2.39). New users were less likely to use psychoactive substances (OR = 0.59, 0.36-0.94). They were also more likely to be concurrent users of psychotherapy (OR = 4.79, 2.87-8.08), but less likely to have started it before the pandemic (OR = 0.14, 0.08-0.25).

**Table. 3.**
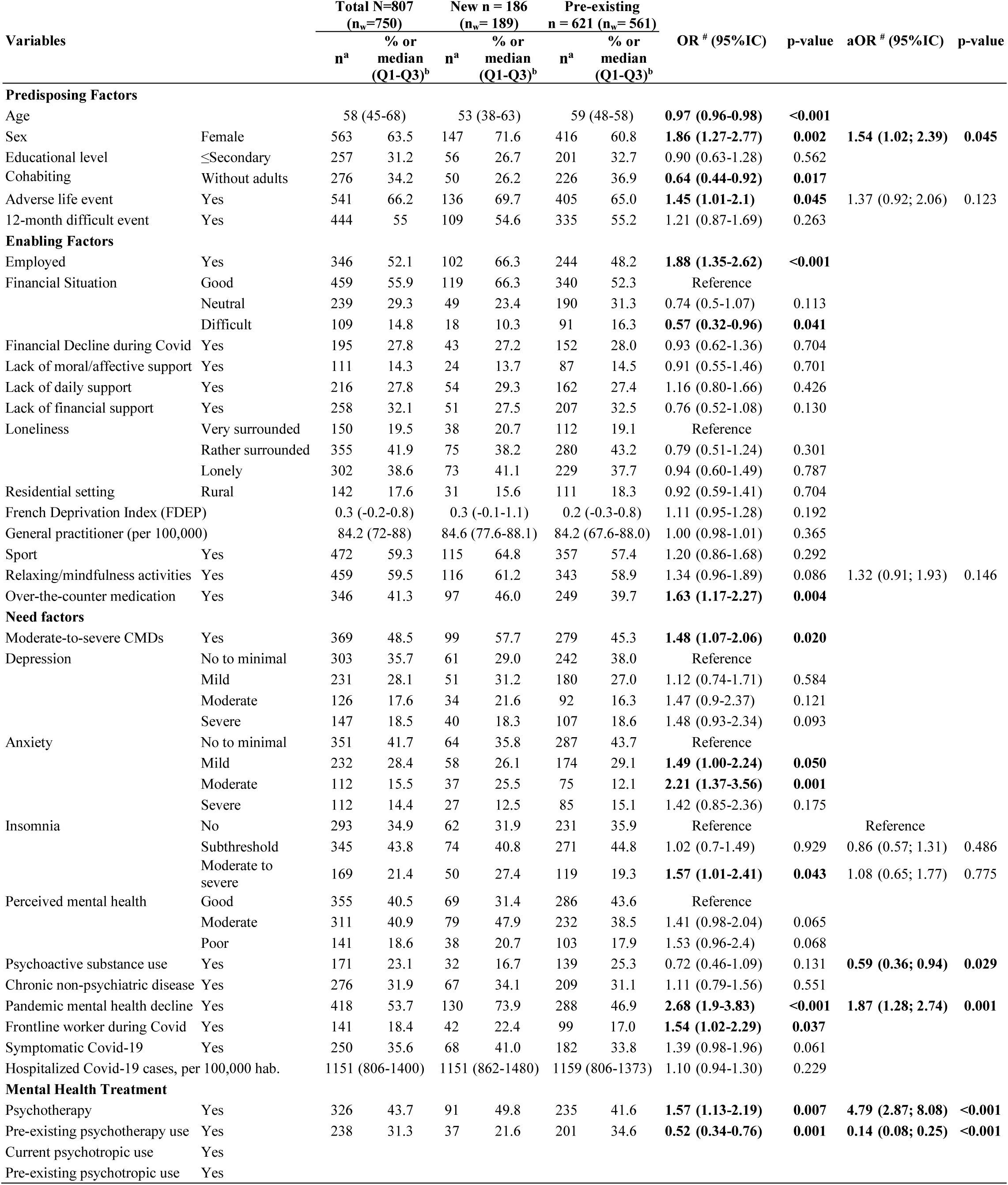
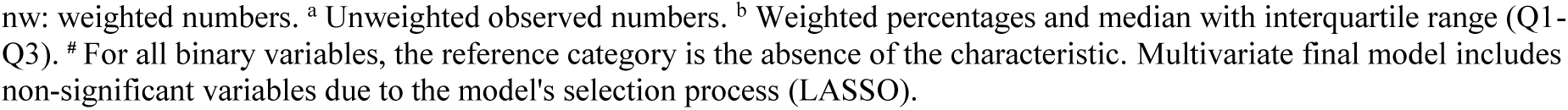
Comparison of new and pre-existing users of pharmacotherapy for common mental disorders during the pandemic.

|  |  | Total N=807<br>(n<sub>w</sub>=750) |  | New n = 186<br>(n<sub>w</sub>= 189) |  | Pre-existing<br>n = 621 (n<sub>w</sub>= 561) |  |  |  |  |  |
| --- | --- | --- | --- | --- | --- | --- | --- | --- | --- | --- | --- |
| Variables |  | n<sup>a</sup> | % or<br>median<br>(Q1-Q3)<sup>b</sup> | n<sup>a</sup> | % or<br>median<br>(Q1-Q3)<sup>b</sup> | n<sup>a</sup> | % or<br>median<br>(Q1-Q3)<sup>b</sup> | OR <sup>#</sup> (95%IC) | p-value | aOR <sup>#</sup> (95%IC) | p-value |
| Predisposing Factors |  |  |  |  |  |  |  |  |  |  |  |
| Age |  | 58 (45-68) |  | 53 (38-63) |  | 59 (48-58) |  | **0.97 (0.96-0.98)** | **<0.001** |  |  |
| Sex | Female | 563 | 63.5 | 147 | 71.6 | 416 | 60.8 | **1.86 (1.27-2.77)** | **0.002** | **1.54 (1.02; 2.39)** | **0.045** |
| Educational level | ≤Secondary | 257 | 31.2 | 56 | 26.7 | 201 | 32.7 | 0.90 (0.63-1.28) | 0.562 |  |  |
| Cohabiting | Without adults | 276 | 34.2 | 50 | 26.2 | 226 | 36.9 | **0.64 (0.44-0.92)** | **0.017** |  |  |
| Adverse life event | Yes | 541 | 66.2 | 136 | 69.7 | 405 | 65.0 | **1.45 (1.01-2.1)** | **0.045** | 1.37 (0.92; 2.06) | 0.123 |
| 12-month difficult event | Yes | 444 | 55 | 109 | 54.6 | 335 | 55.2 | 1.21 (0.87-1.69) | 0.263 |  |  |
| Enabling Factors |  |  |  |  |  |  |  |  |  |  |  |
| Employed | Yes | 346 | 52.1 | 102 | 66.3 | 244 | 48.2 | **1.88 (1.35-2.62)** | **<0.001** |  |  |
| Financial Situation | Good | 459 | 55.9 | 119 | 66.3 | 340 | 52.3 | Reference |  |  |  |
|  | Neutral | 239 | 29.3 | 49 | 23.4 | 190 | 31.3 | 0.74 (0.5-1.07) | 0.113 |  |  |
|  | Difficult | 109 | 14.8 | 18 | 10.3 | 91 | 16.3 | **0.57 (0.32-0.96)** | **0.041** |  |  |
| Financial Decline during Covid | Yes | 195 | 27.8 | 43 | 27.2 | 152 | 28.0 | 0.93 (0.62-1.36) | 0.704 |  |  |
| Lack of moral/affective support | Yes | 111 | 14.3 | 24 | 13.7 | 87 | 14.5 | 0.91 (0.55-1.46) | 0.701 |  |  |
| Lack of daily support | Yes | 216 | 27.8 | 54 | 29.3 | 162 | 27.4 | 1.16 (0.80-1.66) | 0.426 |  |  |
| Lack of financial support | Yes | 258 | 32.1 | 51 | 27.5 | 207 | 32.5 | 0.76 (0.52-1.08) | 0.130 |  |  |
| Loneliness | Very surrounded | 150 | 19.5 | 38 | 20.7 | 112 | 19.1 | Reference |  |  |  |
|  | Rather surrounded | 355 | 41.9 | 75 | 38.2 | 280 | 43.2 | 0.79 (0.51-1.24) | 0.301 |  |  |
|  | Lonely | 302 | 38.6 | 73 | 41.1 | 229 | 37.7 | 0.94 (0.60-1.49) | 0.787 |  |  |
| Residential setting | Rural | 142 | 17.6 | 31 | 15.6 | 111 | 18.3 | 0.92 (0.59-1.41) | 0.704 |  |  |
| French Deprivation Index (FDEP) |  | 0.3 (-0.2-0.8) |  | 0.3 (-0.1-1.1) |  | 0.2 (-0.3-0.8) |  | 1.11 (0.95-1.28) | 0.192 |  |  |
| General practitioner (per 100,000) |  | 84.2 (72-88) |  | 84.6 (77.6-88.1) |  | 84.2 (67.6-88.0) |  | 1.00 (0.98-1.01) | 0.365 |  |  |
| Sport | Yes | 472 | 59.3 | 115 | 64.8 | 357 | 57.4 | 1.20 (0.86-1.68) | 0.292 |  |  |
| Relaxing/mindfulness activities | Yes | 459 | 59.5 | 116 | 61.2 | 343 | 58.9 | 1.34 (0.96-1.89) | 0.086 | 1.32 (0.91; 1.93) | 0.146 |
| Over-the-counter medication | Yes | 346 | 41.3 | 97 | 46.0 | 249 | 39.7 | **1.63 (1.17-2.27)** | **0.004** |  |  |
| Need factors |  |  |  |  |  |  |  |  |  |  |  |
| Moderate-to-severe CMDs | Yes | 369 | 48.5 | 99 | 57.7 | 279 | 45.3 | **1.48 (1.07-2.06)** | **0.020** |  |  |
| Depression | No to minimal | 303 | 35.7 | 61 | 29.0 | 242 | 38.0 | Reference |  |  |  |
|  | Mild | 231 | 28.1 | 51 | 31.2 | 180 | 27.0 | 1.12 (0.74-1.71) | 0.584 |  |  |
|  | Moderate | 126 | 17.6 | 34 | 21.6 | 92 | 16.3 | 1.47 (0.9-2.37) | 0.121 |  |  |
|  | Severe | 147 | 18.5 | 40 | 18.3 | 107 | 18.6 | 1.48 (0.93-2.34) | 0.093 |  |  |
| Anxiety | No to minimal | 351 | 41.7 | 64 | 35.8 | 287 | 43.7 | Reference |  |  |  |
|  | Mild | 232 | 28.4 | 58 | 26.1 | 174 | 29.1 | **1.49 (1.00-2.24)** | **0.050** |  |  |
|  | Moderate | 112 | 15.5 | 37 | 25.5 | 75 | 12.1 | **2.21 (1.37-3.56)** | **0.001** |  |  |
|  | Severe | 112 | 14.4 | 27 | 12.5 | 85 | 15.1 | 1.42 (0.85-2.36) | 0.175 |  |  |
| Insomnia | No | 293 | 34.9 | 62 | 31.9 | 231 | 35.9 | Reference |  | Reference |  |
|  | Subthreshold | 345 | 43.8 | 74 | 40.8 | 271 | 44.8 | 1.02 (0.7-1.49) | 0.929 | 0.86 (0.57; 1.31) | 0.486 |
|  | Moderate to severe | 169 | 21.4 | 50 | 27.4 | 119 | 19.3 | **1.57 (1.01-2.41)** | **0.043** | 1.08 (0.65; 1.77) | 0.775 |
| Perceived mental health | Good | 355 | 40.5 | 69 | 31.4 | 286 | 43.6 | Reference |  |  |  |
|  | Moderate | 311 | 40.9 | 79 | 47.9 | 232 | 38.5 | 1.41 (0.98-2.04) | 0.065 |  |  |
|  | Poor | 141 | 18.6 | 38 | 20.7 | 103 | 17.9 | 1.53 (0.96-2.4) | 0.068 |  |  |
| Psychoactive substance use | Yes | 171 | 23.1 | 32 | 16.7 | 139 | 25.3 | 0.72 (0.46-1.09) | 0.131 | **0.59 (0.36; 0.94)** | **0.029** |
| Chronic non-psychiatric disease | Yes | 276 | 31.9 | 67 | 34.1 | 209 | 31.1 | 1.11 (0.79-1.56) | 0.551 |  |  |
| Pandemic mental health decline | Yes | 418 | 53.7 | 130 | 73.9 | 288 | 46.9 | **2.68 (1.9-3.83)** | **<0.001** | **1.87 (1.28; 2.74)** | **0.001** |
| Frontline worker during Covid | Yes | 141 | 18.4 | 42 | 22.4 | 99 | 17.0 | **1.54 (1.02-2.29)** | **0.037** |  |  |
| Symptomatic Covid-19 | Yes | 250 | 35.6 | 68 | 41.0 | 182 | 33.8 | 1.39 (0.98-1.96) | 0.061 |  |  |
| Hospitalized Covid-19 cases, per 100,000 hab. |  | 1151 (806-1400) |  | 1151 (862-1480) |  | 1159 (806-1373) |  | 1.10 (0.94-1.30) | 0.229 |  |  |
| Mental Health Treatment |  |  |  |  |  |  |  |  |  |  |  |
| Psychotherapy | Yes | 326 | 43.7 | 91 | 49.8 | 235 | 41.6 | **1.57 (1.13-2.19)** | **0.007** | **4.79 (2.87; 8.08)** | **<0.001** |
| Pre-existing psychotherapy use | Yes | 238 | 31.3 | 37 | 21.6 | 201 | 34.6 | **0.52 (0.34-0.76)** | **0.001** | **0.14 (0.08; 0.25)** | **<0.001** |
| Current psychotropic use | Yes |  |  |  |  |  |  |  |  |  |  |
| Pre-existing psychotropic use | Yes |  |  |  |  |  |  |  |  |  |  |

nw: weighted numbers. <sup>a</sup> Unweighted observed numbers. <sup>b</sup> Weighted percentages and median with interquartile range (Q1-Q3). <sup>#</sup> For all binary variables, the reference category is the absence of the characteristic. Multivariate final model includes non-significant variables due to the model's selection process (LASSO).

## 4. Discussion

This study extends existing literature by reporting that one-quarter of the French general population was receiving treatment for CMD two years after the onset of the COVID-19 pandemic, with details on both pharmacotherapy (18%) and psychotherapy (16%) use. Our findings reveal differences between new and pre-existing users of psychotherapy and pharmacotherapy in need-related factors, as well as in enabling factors for psychotherapy users and in predisposing factors for pharmacotherapy users.

Our reported treatment prevalence (26.1%) is consistent with 2021 data from the United States, where a quarter of adults reported receiving prescription medication or therapy for anxiety or depression disorders in the past 4 weeks [39]. Direct comparisons with other studies remain challenging due to differences in outcome definitions and assessment periods. For example, our estimate is higher than the 13.2% of participants in a Canadian survey who reported using mental health services in 2022 [18]. This lower prevalence likely reflects a narrower definition of “mental health service use,” which may not have captured all treatments delivered - such as prescription renewals or general mental health support provided by general practitioners. One in four people receiving treatment for CMD echoes the high mental health burden in high-income countries in Europe, where about 25% report mental health problems [40]. However, this does not reflect treatment coverage among those meeting diagnostic criteria. Many individuals with psychological distress or CMD symptoms still report unmet treatment needs, indicating that access, adequacy, or targeting of treatment remains insufficient [12–16, 18]. Further research is needed to better understand the adequacy between treatments, CMD symptoms, their functioning impact, and patient preferences.

When looking at treatment by therapy types, our results for pharmacotherapy align with trends observed in several countries [39, 41, 42]. We also found that 16% of French adults receive psychotherapy for CMDs, similar to the 18% of adults who consulted their GP and 17% who saw mental health specialists during the pandemic, according to another French survey [43]. This represents a significant increase compared to the 7% reported in 2005 across four regions in France, likely reflecting higher needs, evolving clinical recommendations, efforts to reduce the stigma surrounding mental health care, and changes in reimbursement policies for psychotherapy [1, 2, 42, 44]. However, our findings of similar use of psychotherapy (16%) and pharmacotherapy (18%) suggest a divergence between clinical practices and guidelines, which typically recommend psychotherapy as a first-line treatment [23, 45–48]. This trend was less pronounced in new users than in those with pre-existing ones, potentially due to factors such as different disease stages, or evolving societal attitudes toward mental health. Notably, one-third of new users received psychotropic treatment alone, while combined psychotherapy and pharmacotherapy has been reported as the optimal treatment of CMDs [49–52]. These discrepancies may be attributed to systemic barriers, such as limited insurance coverage for psychotherapy, patient’s or clinicians’ preference for pharmacotherapy, or reduced access to psychotherapy early in the pandemic, when teleconsultations were less widespread.

New users of psychotherapy differed from pre-existing users in pandemic-related need factors and reported fewer financial difficulties. The significant association between symptomatic COVID-19 and psychotherapy use is consistent with studies showing that symptomatic infectious disease responses can heighten psychological distress [53, 54]. Fewer financial difficulties among new users align with evidence that individuals with lower income were more likely to face barriers to mental health care during the pandemic [18]. Previous research also indicates that regions with higher socioeconomic resources had greater mental health service use, while CMDs disproportionately affected disadvantaged populations, and that financial hardship was strongly associated with unmet mental health needs [10, 17, 18]. Income loss among pre-existing users since their first treatment may also explain our findings, as depression increases the risk of unemployment or reduced income [55]. However, our cross-sectional design does not allow us to determine causality or temporal relationships. Given that this difference was observed only for psychotherapy, which often requires out-of-pocket payment or is less frequently reimbursed compared to pharmacotherapy, and considering existing literature, these results likely reflect, at least partially, inequities in access to psychotherapy. Finally, the lack of significance for predisposing factors in multivariate analyses suggests that new use of psychotherapy was primarily driven by greater need and structural factors, rather than by a shift in help-seeking behavior.

As expected, new users of pharmacotherapy were also characterized by an increased perceived mental health decline. Lower psychoactive substance use among new users may reflect differences in the populations accessing care, with new users potentially having fewer pre-existing substance use issues. The sustained association between female sex and new pharmacotherapy use is consistent with pre-pandemic research indicating that women are more likely than men to use anxiolytics for managing acute social and emotional issues [56]. The lack of significance for enabling factors in multivariate analyses showed that structural barriers are less pronounced for pharmacotherapy compared to psychotherapy, probably due to better insurance coverage and better care provision.

A key strength of this study is the availability of data on both psychotherapy and pharmacotherapy for CMD, as well as on predisposing, enabling, and need factors from a national survey. In addition, this study, conducted in a crisis context, offers exploratory insights that could help guide understanding of mental health service use in similar situations in the future. However, several limitations should be noted. First, despite the weighting, the voluntary and non-random nature of the sample may limit generalizability. Second, the cross-sectional design and reliance on self-reported data for treatment initiation, along with the absence of pre-pandemic baseline data, may introduce recall bias and do not allow us to distinguish whether observed differences are due to actual group differences or to temporal effects. Third, our analysis included only individuals receiving treatment at the time of the survey, which may underestimate the total number of people who initiated treatment during the pandemic. Fourth, detailed information on treatment type or provider (e.g., psychologist, psychiatrist, general practitioner) was not available. Finally, there may still be residual confounding. Finally, we used LASSO regression without a priori variable grouping to allow the model to identify the most discriminant factors among correlated variables, which aligns with our exploratory objective of comparing profiles rather than estimating isolated causal effects.

In conclusion, this study shows that one in four French adults was receiving treatment for CMD two years after the onset of the COVID-19 pandemic, prevailing the mental health burden. New treatment-seeking was primarily driven by pandemic-related stressors, while financial disparities likely influence access to psychotherapy. Future research should assess remaining unmet needs and the long-term trends of these patterns. Policy efforts should prioritize equitable access to evidence-based mental healthcare, in particular to psychotherapy.

## Data Availability

All data produced in the present study are available upon reasonable request to the authors

## List of abbreviations

COVID-19: Coronavirus Disease 2019
CMD: Common Mental Disorders
PHQ-9: Patient Health Questionnaire-9
GAD-7: Generalized Anxiety Disorder-7
ISI: Insomnia Severity Index
ATC: Anatomical Therapeutic Chemical classification system
OTC: Over-the-Counter
CCTIRS: Comité Consultatif sur le Traitement de l’Information en matière de Recherche dans le domaine de la Santé - French Advisory Committee on Information Processing in Health Research
CNIL: Commission Nationale de l’Informatique et des Libertés - French Data Protection Authority
INSEE: Institut National de la Statistique et des Études Économiques - French National Institute of Statistics and Economic Studies
LASSO: Least Absolute Shrinkage and Selection Operator
CI: Confidence Interval

## Declarations

### Consent for publication

Not applicable.

### Availability of data and materials

The datasets used and/or analysed during the current study are available from the corresponding author on reasonable request.

### Authors’ contributions

All authors participated in the design of the study. MP conducted the analyses. MP wrote the first draft of this report. All authors contributed to the review of the manuscript, read the manuscript and approved the final version.

